# A retrospective observational study to investigate the effect of frailty on outcomes of older adults admitted with major trauma

**DOI:** 10.1101/2021.12.29.21268277

**Authors:** Philip Braude, Omar Bouamra, Frances Parry, Fiona Lecky, Ben Carter

**Affiliations:** CLARITY (Collaborative Ageing Research) group, North Bristol Trust; University of Manchester; University of Sheffield, TARN Research Director; King’s Clinical Trials Unit

**Keywords:** Frailty, trauma, elderly care, geriatric medicine

## Abstract

A protocol for a retrospective observational study to investigate the effect of frailty on outcomes of older adults admitted with major trauma. Patient data will be examined held within the Trauma and Audit Research Network (TARN) database, from 23 Major Trauma Centres in England, between April 2019 and March 2020. The two research questions are: 1) Is there any association between frailty and clinical outcomes in older people admitted with serious traumatic injuries? 2) Is there any association between a geriatrician review and clinical outcomes in an older people admitted with serious traumatic injuries? Patients 65 years old or over will be included. The primary outcome will be mortality as measured by time from hospital admission to death. Secondary outcomes will include day 30 mortality, post injury complications, and length of hospital stay. A mixed-effects multivariable Cox proportional hazards model will be used to analyse the data. A statistical analysis plan has been produced separately for each research question.

**Signature Page:** The undersigned confirm that the following protocol has been agreed and accepted and that the Chief Investigator agrees to conduct the study in compliance with the approved protocol and will adhere to the principles outlined in the Declaration of Helsinki, the Sponsor’s SOPs, and other regulatory requirement.

I agree to ensure that the confidential information contained in this document will not be used for any other purpose other than the evaluation or conduct of the investigation without the prior written consent of the Sponsor

I also confirm that I will make the findings of the study publically available through publication or other dissemination tools without any unnecessary delay and that an honest accurate and transparent account of the study will be given; and that any discrepancies from the study as planned in this protocol will be explained.

**For and on behalf of the Study Sponsor:** 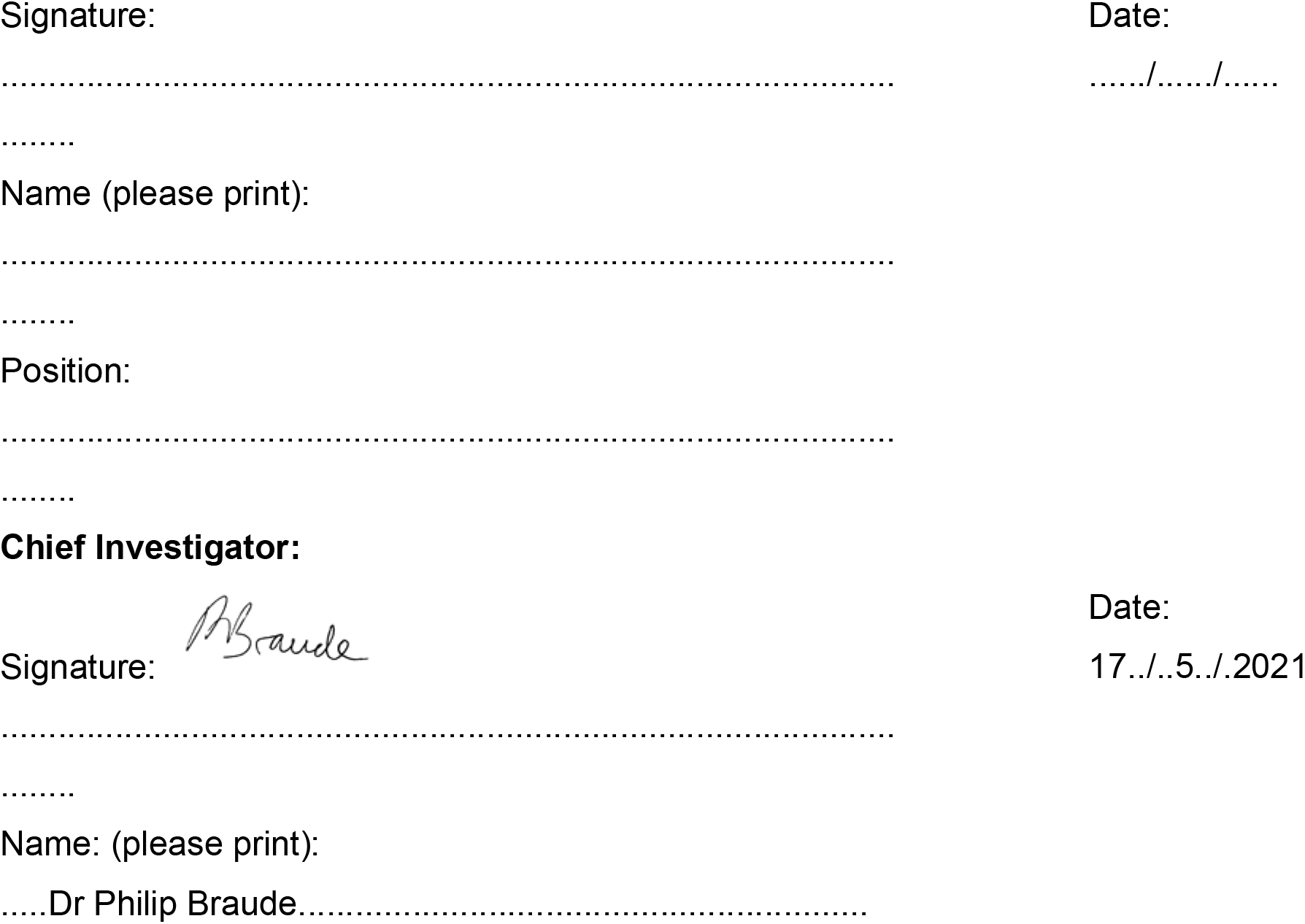

*Key Study Contacts:* 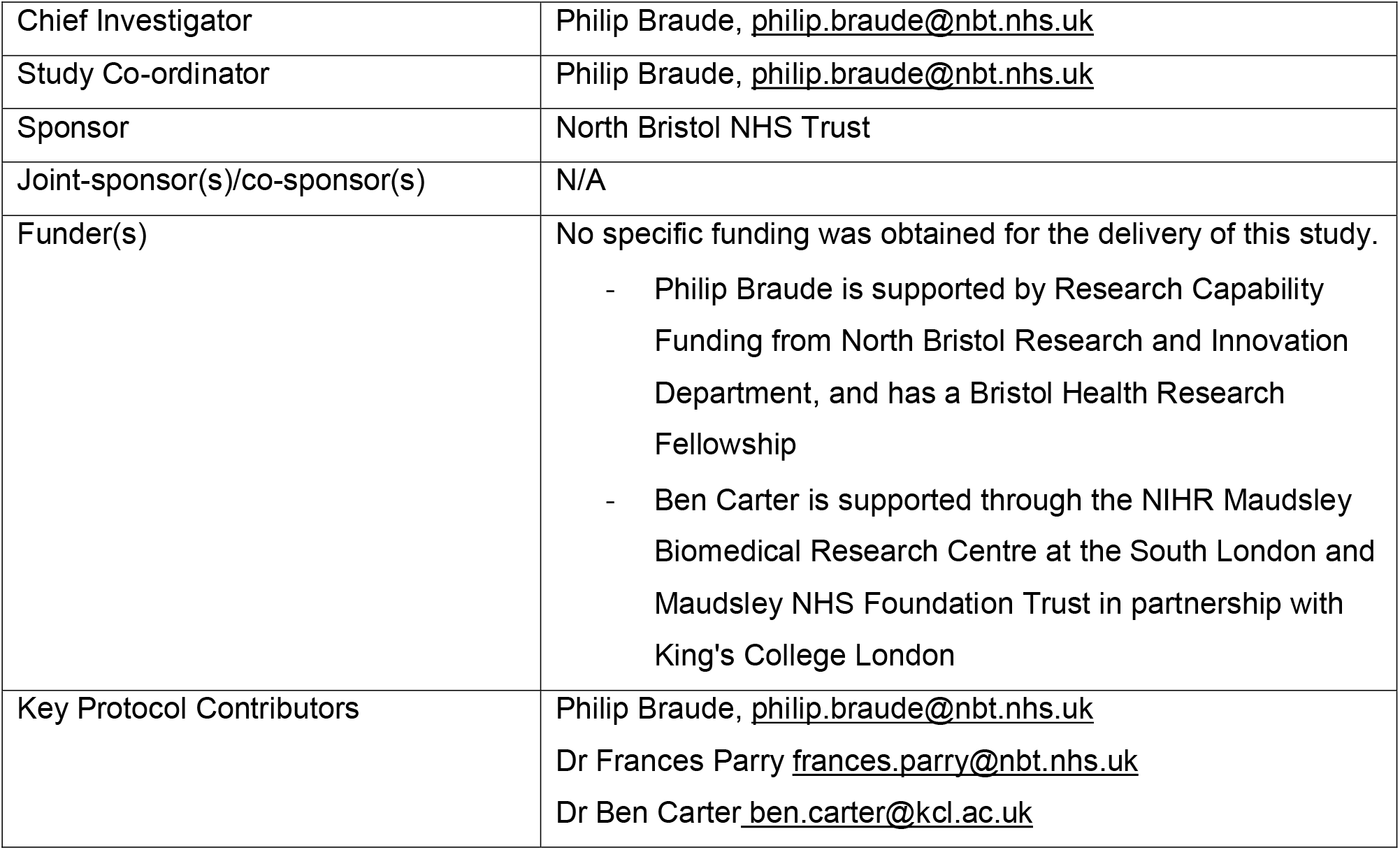

*Study Summary:* 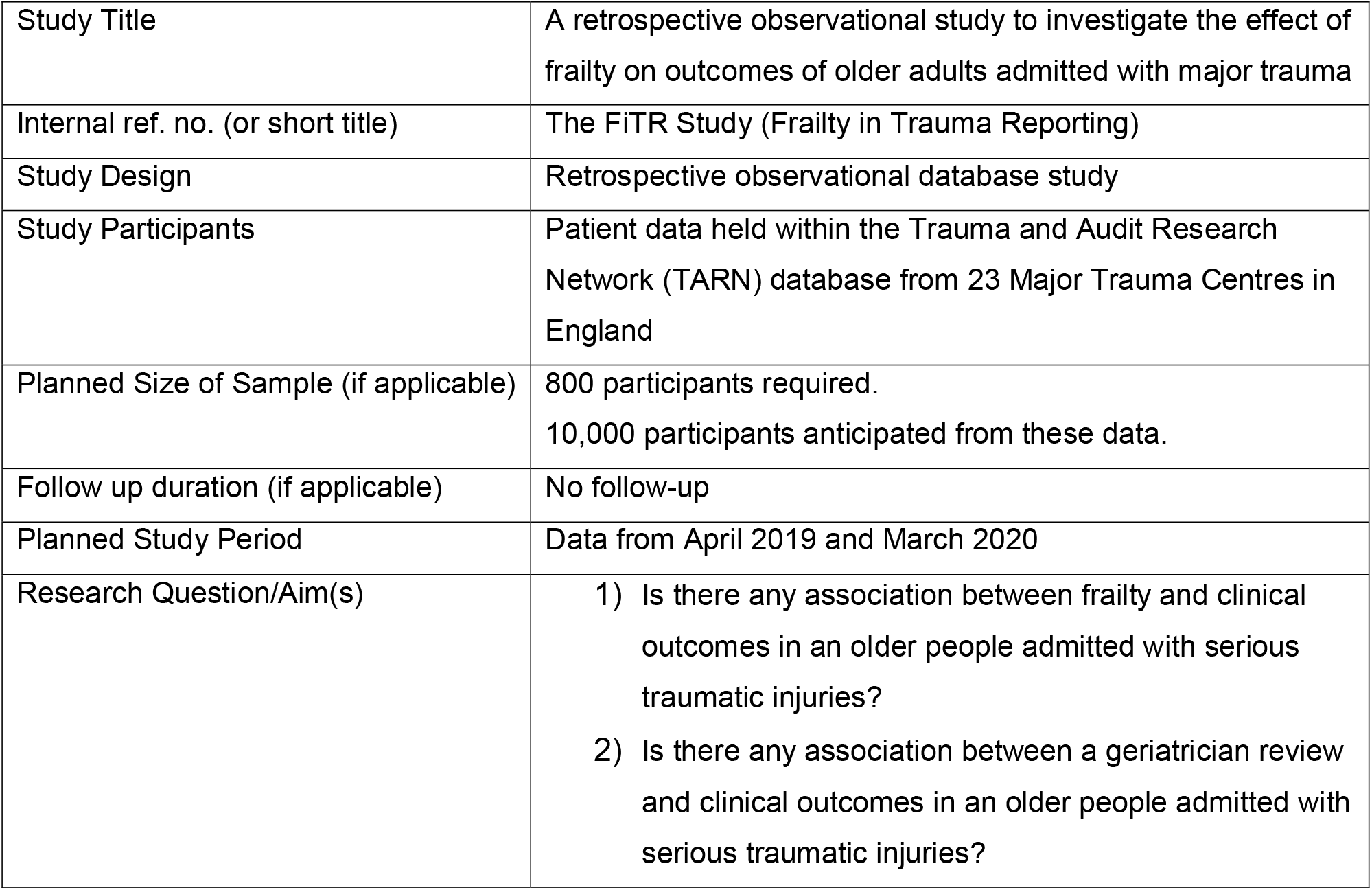

*Plain Language Summary:* The majority of major injuries admitted to hospital are now in older people, with a fall from standing height being the most common reason for injury. Our study will look at older adults admitted to hospital with serious traumatic injuries across England and Wales. It will aim to work out firstly if there is an effect of frailty on peoples’ survival after injury. Frailty is the reserve a person has to cope with illness and is a measure of a person slowing down over time usually due to the collection of lots of health problems. Secondly, we will look at if being seen by an old age specialist (geriatrician) has an effect on a person’s chances of surviving their injuries. The records we will look at are held collected routinely and held by a national database run by the Trauma and Audit Research Network (TARN) from the University of Manchester. They collect information from the 23 centres of major trauma excellence around the country and help researchers to work with them to access the anonymous data for specific research questions. We aim to use these results to help clinicians and health systems to improve how the fund and deliver care for older people.

*Funding and Support in Kind:* 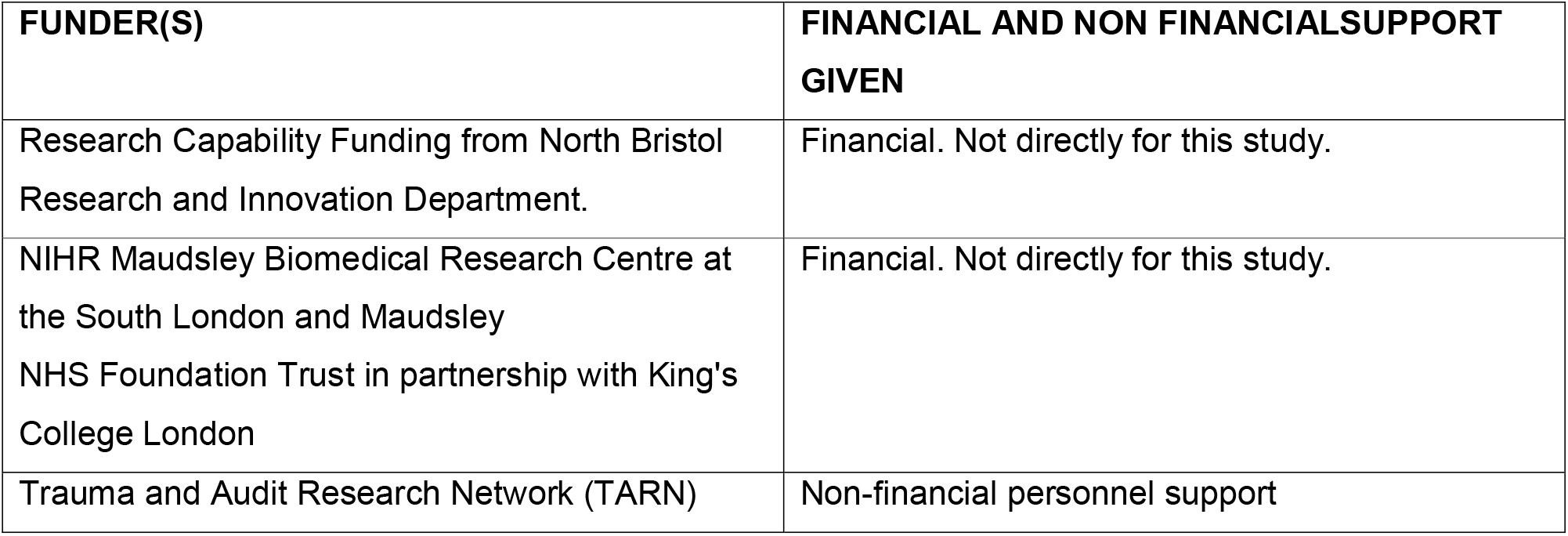

*Role of Study Sponsor and Funder:* North Bristol Trust R&I - supporting the development of this work. No direct influence in any part of the work. TARN – supporting development of the work. Will have input into the development, analysis and write-up of the work.

*Study Steering Group:* 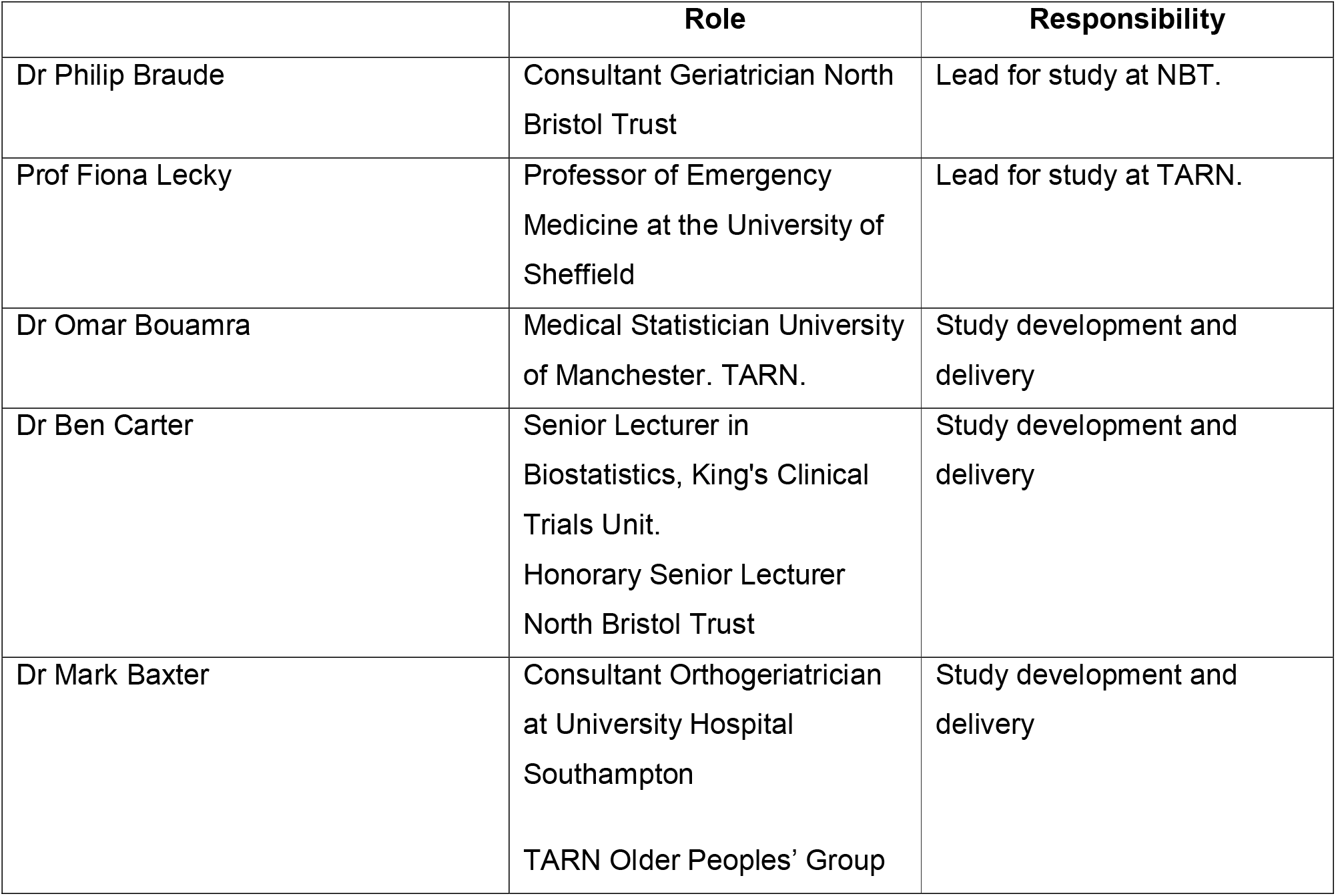

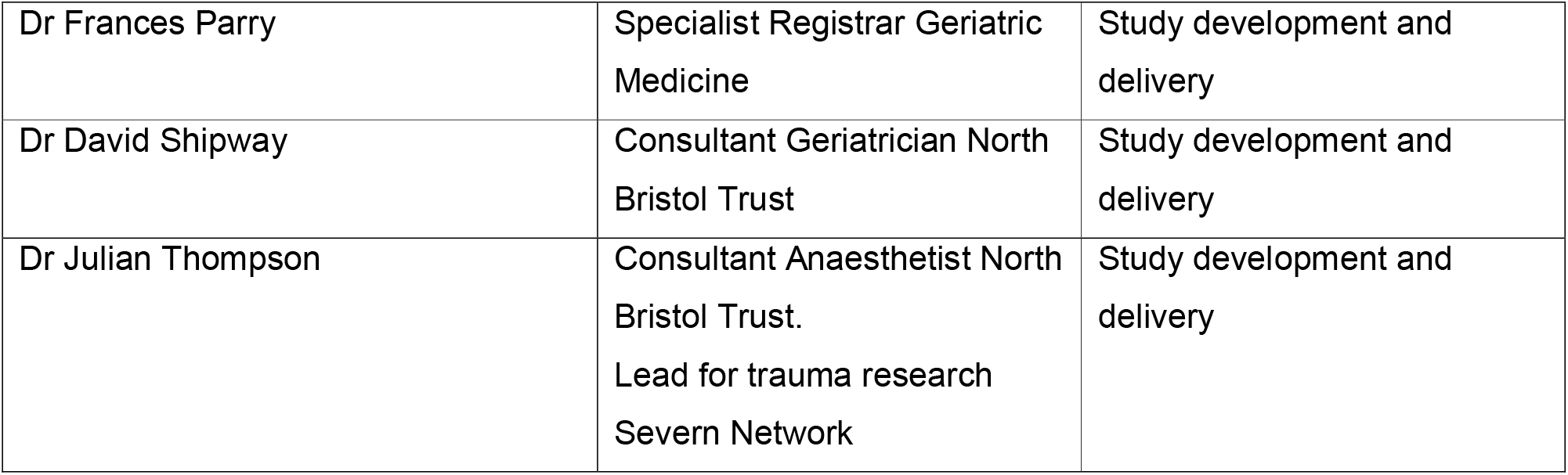

*Protocol Contributors:* 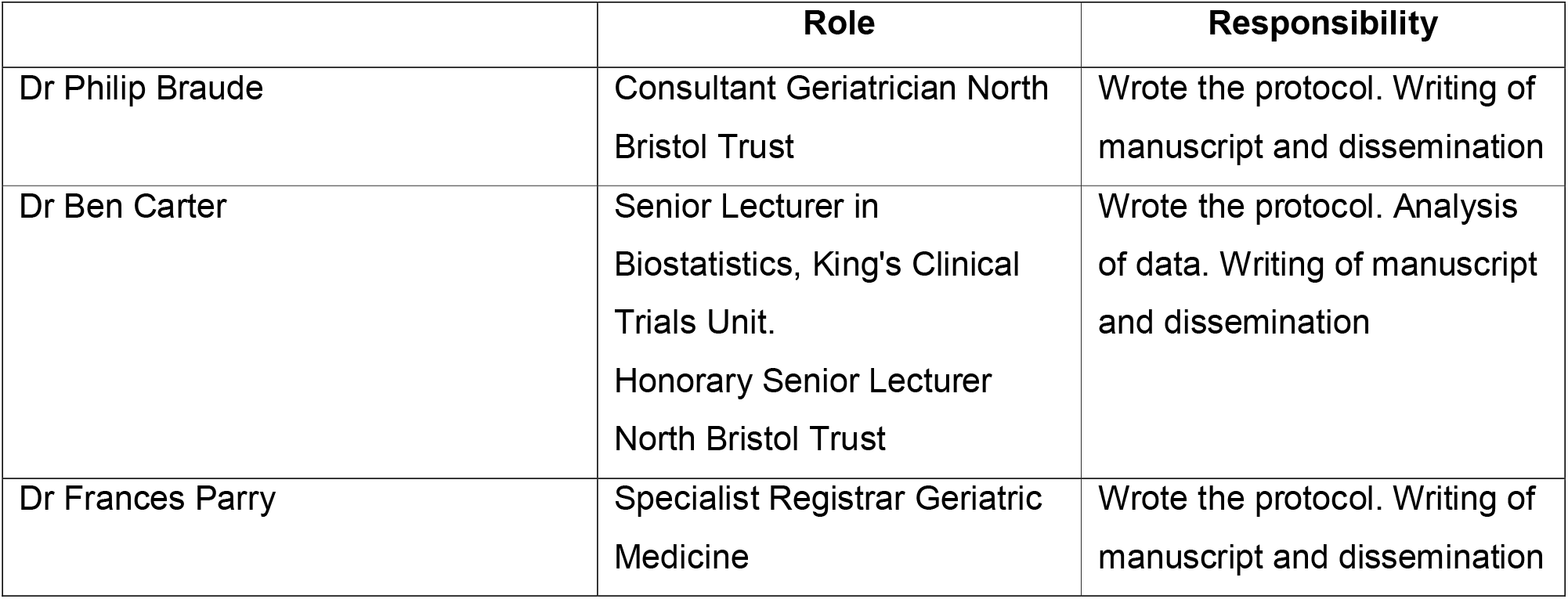 No patients were involved in the development of the protocol.

*Study Flow Chart:* 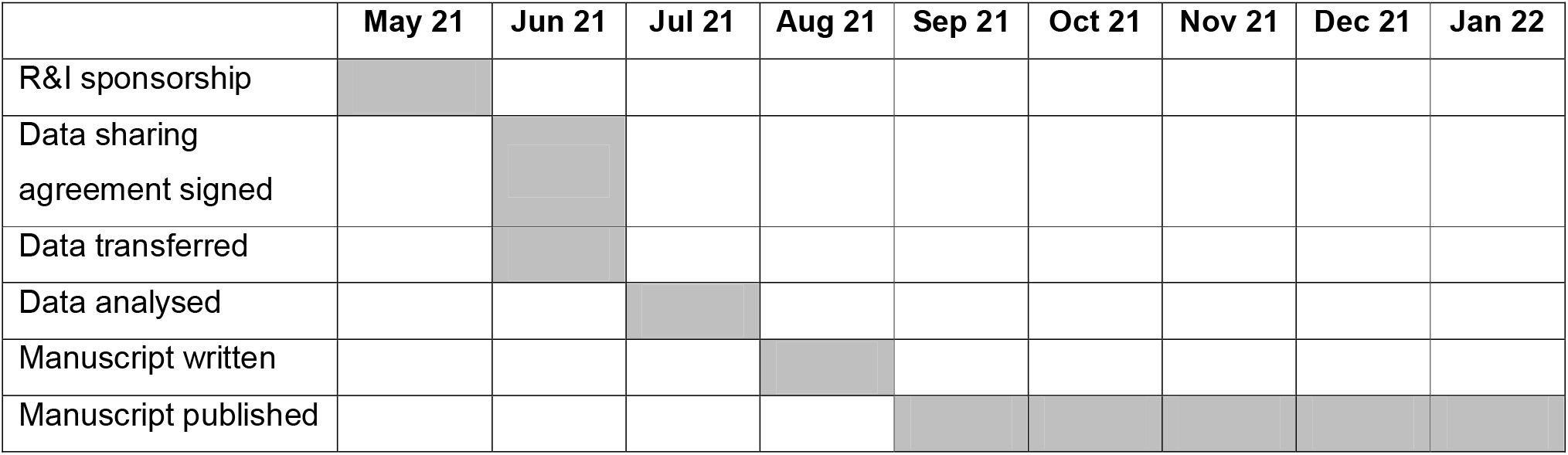

*Study Protocol:* A retrospective observational study to investigate the effect of frailty on outcomes of older adults admitted with major trauma.

## 1 Background

Over half of patients sustaining major trauma are aged 65 and older. They experience a higher rate of mortality compared to younger patients, even at matched injury severity scores (ISS) [1]. In 2019, in recognition of this, NHS England and NHS Improvement mandated as part of the best practice tariff (BPT) that a frailty assessment be performed by a specialist trainee, consultant or non-consultant grade in geriatric medicine within 72 hours of admission [2]. The Trauma and Audit Research Network (TARN) database collates data across 23 MTCs in England. Patients’ frailty scores are recorded on the TARN database, alongside their injury severity score, mechanism of injury, and various outcome measures.

The frailty assessment uses the Clinical Frailty Scale (CFS), which has been validated as a predictor of institutionalisation and mortality in the non-trauma healthcare setting [3]. There have been two studies in North America showing an association between CFS and outcome in major trauma patients [4, 5]. The only UK study, undertaken at North Bristol Trust, examined the outcomes after 363 patients

≥65 years old admitted to the MTC had a CFS assigned and followed over a 5 month period [6]. They found that CFS is an independent predictor of 30-day mortality, inpatient delirium and increased level of care at discharge. These data were underpowered to detect an effect of CFS on length of hospital stay.

In a similar work looking at frailty and patient outcomes, the ELF study (Emergency Laparotomy and Frailty) [7] found that the CFS is a predictor of mortality and level of care required at discharge, and that it should be used to inform decision making and discharge planning in surgical units. Beyond this, the National Emergency Laparotomy Audit (NELA) published a report demonstrating that postoperative review by a geriatrician was associated with lower mortality [8].

We aim to look at similar outcome measures as both analysed by Rickard et al, and within NELA.

## 2 Rationale

Understanding the effect of frailty on outcomes will help with service development both at the local level and with regard the national trauma strategy. Having clarify on the effects of frailty can guide early patient discussions about outcome after traumatic injury, provide information for service resource planning including pathways incorporating frailty, and support mainstream local funding of geriatric services – rather than reliance on BPT.

## 3 Theoretical Framework

This is an observational database study to explore the effects of frailty on outcomes. TARN database is the only repository of frailty scores in the trauma population. Although the Best Practice Tariff encourages scoring of the CFS for all older people, this is not mandatory, and not routinely held in any other database.

No other study has examined the CFS scored on a national level to compare. A retrospective approach is being taken in order to explore the completion rates since introduction.

## 4 Research Question/Aims

To investigate the associated outcomes of major trauma in older patients with frailty using the TARN database.

### 4.1 Objectives

#### Objective 1

Is there any association between frailty and clinical outcomes in an older trauma cohort?

#### Objective 2

Is there any effect on the outcomes of an older trauma cohort when reviewed by a geriatrician?

### 4.2 Outcome

Primary outcome:

- Mortality (time from hospital admission to death) Secondary outcomes:
- day 30 mortality
- post injury complications
- length of hospital stay

## 5 Study Design, Methods of Data Collection and Data Analysis

Retrospective observational cohort study using the TARN database from April 2019 to March 2020. Major trauma centres will be de-identified using a numerical key (MTC 1 to 23). No patient identifiers will be used. Data will be anonymously transferred to North Bristol Trust for analysis. Statistical methods will have been developed with a statistician approved by TARN. The study will be written in accordance with the STROBE statement [10].

### Objective 1

Patients will be analysed within each CFS category separately (1 to 9) and trichotomised as: non-frail, mild to moderate frailty, and severely frail (CFS 1–4 vs 5-6 vs 7-9). Time to mortality and time to discharge outcomes will be analysed with a mixed-effects multivariable Cox proportional hazards models, with a random effect for site to account for variation across site, and adjusted for patient age group, CFS, ageless-CCI, ISS and surgery (yes or no). Binary outcomes (Day 30 mortality, major complication) will be analysed using a mixed effects logistic regression, in a format consistent with the primary outcome, reported with an odds ratio (OR). Data will be reported as both crude and adjusted OR or HR respectively; with associated 95% confidence intervals. Each time-to-event analysis will be accompanied by a Kaplan-Meier survival plot, with accompanied at risk table. Breaches of baseline proportionality will be assessed using log-log plots. Missing data will be explored prior to analysis, and sensitivity analysis will be conducted to test key assumptions.

### Objective 2

Compare outcomes of patients with a CFS (as surrogate for review by a geriatrician) and patients with missing CFS (as a surrogate of not having been reviewed). Univariate analysis will be performed to assess between patient outcome those who have had a CFS, and those who have not. Analysis will be carried out consistent with Objective 1.

## 6 Study Setting

The Trauma Audit and Research Network (TARN) is run by the University of Manchester The database collects and processes data on moderately and severely injured patients in England and Wales. Data is inputted by each trauma centre to allow benchmarking between centres.

Only the 23 adult Major Trauma Centres have been collecting and submitted frailty scores. Only these 23 Trusts will be examined in the study. Data will be transferred from TARN via data sharing agreement to North Bristol Trust.

## 7 Sample and Recruitment

### 7.1 Eligibility Criteria

#### 7.1.1 Inclusion criteria

Objective 1 and 2:

- Patients ≥ 65 years old
- Eligibility for inclusion on TARN database:
  - Trauma patient (excluding military personnel and iatrogenic injuries)
  - LOS ≥3 days/admitted to critical care (irrespective of LOS)/death
  - Patients with specific isolated injuries
- Admitted to the MTC

#### 7.1.2 Exclusion criteria

Objective 1:

- No CFS recorded in database
- Patients younger than 65 Objective 2:
- Patients younger than 65

### 7.2 Sampling

#### 7.2.1 Size of Sample

From a single MTC study we found that one year all-cause mortality was approximately 10% within CFS 1-3, and this increased to 20% in CFS 4-6. In order to detect this difference (Hazard Ratio = 2.12) with a 2.5% significance level (adjusted for two comparisons), and 90% power 100 events would be needed from CFS 1-6. We estimate that no more than 10% of patients will censored due to loss to follow up so will require a minimum of 800 participants and this will be inflated to 1000 by including CFS 1-9.

Using NBT data we included approximately 500 participants in TARN as a typical MTC. Thus, it is anticipated that from 20 MTC would provide approximately 10,000 participants and be more than adequately powered to carry to address this research question.

#### 7.2.2 Sampling Technique

Convenience sampling will be used. All patient level data that has been submitted to TARN and meets the inclusion criteria will be analysed.

## 8 Ethical and Regulatory Considerations

### 8.1 Assessment and management of risk

1. Data sharing agreement not completed: the study will not proceed
2. Data security issues: Data will be stored on Trust computer systems with password protection. Only those in the study team will have access to these data.
3. Identifiable patient information issues: Analysis will take place using anonymised records. Patient level and hospital level identifiers will be removed.

### 8.2 Research Ethics Committee (REC) and other Regulatory review

Approval for the study was sought from the Human Research Authority via the Integrated Research Application System (IRAS number: 303960). HRA approval was gained on 12^th^ November 2021 with Research Ethic Committee reference 21/HRA/3717.

#### Regulatory Review & Compliance

- Before any site can enrol patients into the study, the Chief Investigator/Principal Investigator or designee will ensure that appropriate approvals from participating organisations are in place. Specific arrangements on how to gain approval from participating organisations are in place and comply with the relevant guidance. Different arrangements for NHS and non NHS sites are described as <u>relevant</u>.
- For any amendment to the study, the Chief Investigator or designee, in agreement with the sponsor will submit information to the appropriate body in order for them to issue approval for the amendment. The Chief Investigator or designee will work with sites (R&D departments at NHS sites as well as the study delivery team) so they can put the necessary arrangements in place to implement the amendment to confirm their support for the study as amended.

#### Amendments

The Chief Investigator will liaise with the study sponsor (NBT R&I) to discuss any proposed amendments. If the sponsor wishes to make a substantial amendment the specialist review body (Confidentiality Advisory Group (CAG)) will need to be notified in case the amendment affects their opinion of the study.

Amendments will be tracked with appropriate version controlled protocol changes and stored on Trust computer systems.

### 8.3 Peer review

The protocol has been reviewed by the Chief Investigator and study coordinators prior to being sent for assessment by the nominated Clinical Trials Officer in the Research and Innovation Department at NBT.

### 8.4 Patient & Public Involvement

Public and patient involvement will occur after analysis of the data to ensure appropriate dissemination of the end message. TARN has patient and public involvement on the TARN Board which has oversight of the research portfolio.

### 8.5 Protocol compliance

- Accidental protocol deviations can happen at any time. They will be adequately documented on the relevant forms and reported to the Chief Investigator and Sponsor immediately.
- Deviations from the protocol which are found to frequently recur are not acceptable, will require immediate action and could potentially be classified as a serious breach.

### 8.6 Data protection and patient confidentiality

All investigators and study site staff will comply with the requirements of the Data Protection Act 1998 with regards to the collection, storage, processing and disclosure of personal information and will uphold the Act’s core principles.

Data will be transferred from the University of Manchester (UoM). A data transfer agreement will be signed between NBT R&I and UoM. These data will be identifiable as proprietary to the University of Manchester.

These data will be the responsibility of the Chief / Principal Investigator. Data will be stored on Trust computer systems with password protected folders. Access to the data will be kept to a minimum and only those in the study team. Analysis will take place using anonymised records. Patient level and hospital level identifiers will be removed.

Data will be held until 14-12-23 or until the per permitted research has been completed whichever is sooner. Data is stored in accordance with GDPR and The Data Protection Act 018

### 8.7 Indemnity

This is an NHS-sponsored research study. For NHS sponsored research HSG(96)48 reference no.2 refers. If there is negligent harm during the clinical trial when the NHS body owes a duty of care to the person harmed, NHS indemnity covers NHS staff, medical academic staff with honorary contracts, and those conducting the trial. NHS indemnity does not offer no-fault compensation and is unable to agree in advance to pay compensation for non-negligent harm

### 8.8 Access to the final study dataset

The final dataset will only be accessed by the study team completing the analysis.

## 9 Dissemination Policy

### 9.1 Dissemination policy

The work will be disseminated via peer review journal submission, conference presentation, social media, and press release with embargo date.

### 9.2 Authorship eligibility guidelines and any intended use of professional writers

Authorship will be decided between the study team based on contribution to the final work. No professional writers will be used.

## Supporting information

FiTR 1 Statistical Analysis Plan 0.53

FiTR 2 Statistical Analysis Plan 0.53

FiTR Protocol 1.0 (superceded)

## Data Availability

All data produced in the present study are available upon reasonable request to the authors.

## 11 Appendicies

### 11.1 Appendix 1-Required documentation

Curriculum vitaes of the research team will be submitted

### 11.2 Appendix 2 – Schedule of Procedures

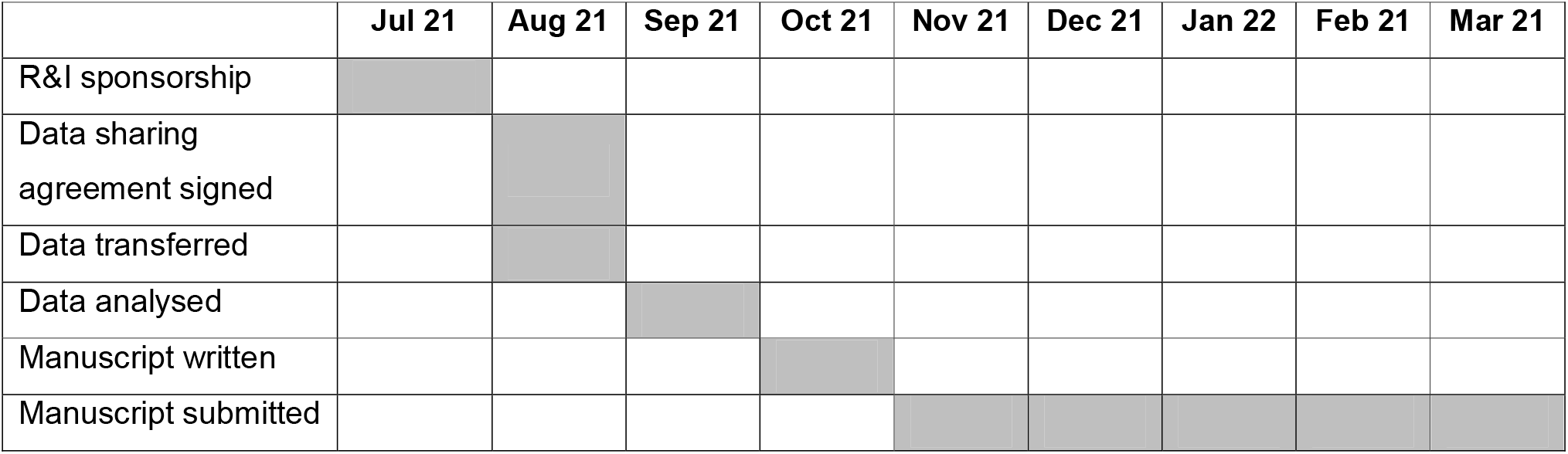

### 11.3 Appendix 3 – Amendment History

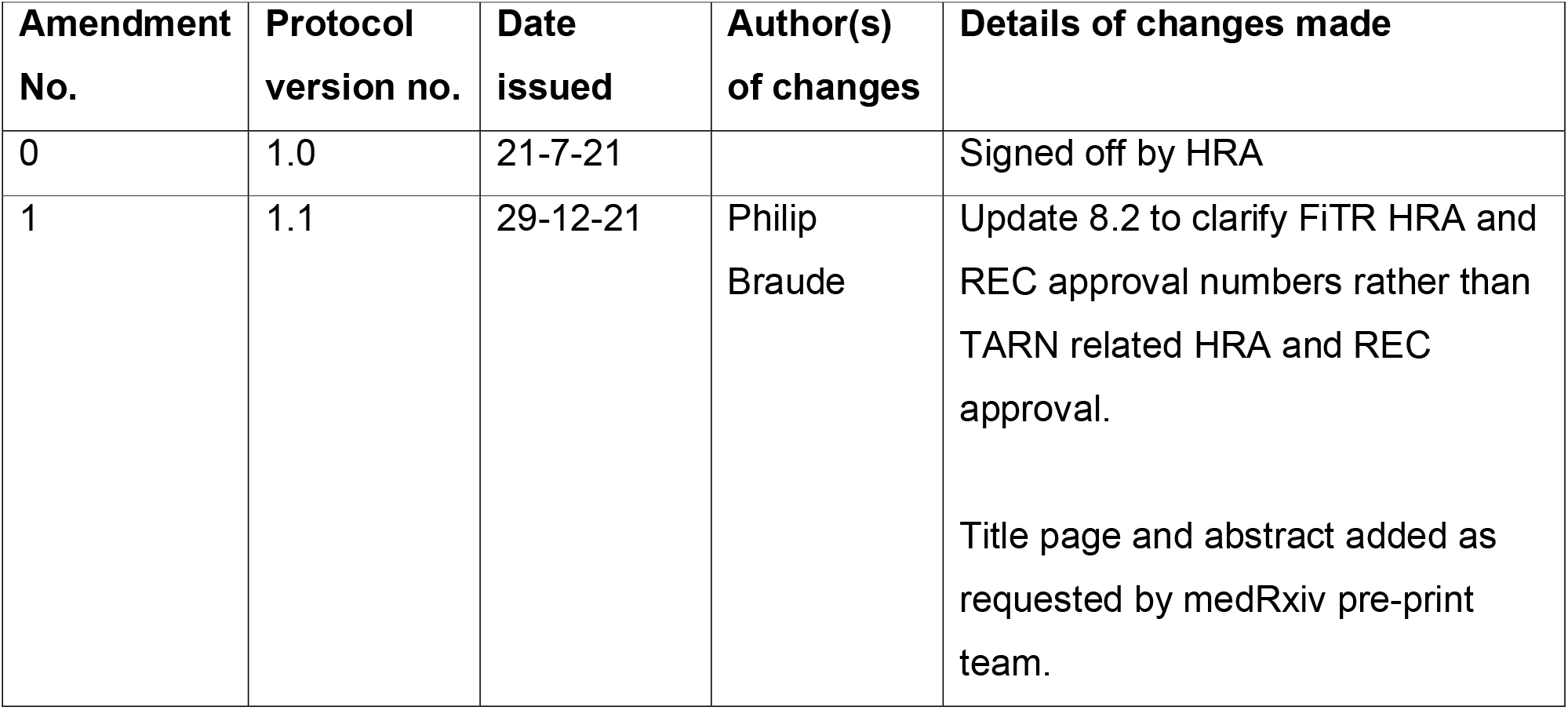

