## Supplementary material for "A retrospective observational study to investigate the effect of frailty on outcomes of older adults admitted with major trauma": FiTR 1 Statistical Analysis Plan 0.53

**Short Title:** The FiTR 1 Study (Frailty in Trauma Reporting)

Statistical Analysis Plan
23rd December, 2021
Version 0.53
IRAS: 303960

***APPROVED***
***By Philip Braude at 11:29 pm, Dec 23, 2021***

**Approved by:**

| | | |
|---|---|---|
| FiTR Study Statistician<br>Dr Ben Carter | Reader in Medical Statistics | 16th December 2021 |
| FiTR Chief Investigator<br>Dr Philip Braude | Consultant Geriatrician | 16th December 2021 |
| TARN Lead Methodologist<br>Dr Omar Bouamra | Medical Statistician | 17th December 2021 |
| TARN Research Director<br>Prof Fiona Lecky | TARN Research Lead | 23rd December 2021 |

**Authors:**

**Study Statistician**

Dr Ben Carter,

Reader in Medical Statistics,

Department of Biostatistics and Health Informatics

CLARITY (Collaborative Ageing Research) group

Institute of Psychiatry, Psychology and Neuroscience

King's College London

**Chief Investigator**

Dr Philip Braude,

Consultant Geriatrician,

CLARITY (Collaborative Ageing Research) group

REACH (Research in Emergency Care Avon Collaborative Hub)

North Bristol NHS Trust

On behalf of FiTR investigators

### Plain Language Summary

FiTR 1 study will look at older adults admitted to hospital with serious traumatic injuries across England. We plan to look at how frailty affects a person's recovery after a major injury. Frailty is the level of physical and mental vulnerability a person has to illness or injury.

The information used for this study is held by the national database run by the Trauma and Audit Research Network (TARN) and is managed by the University of Manchester. TARN collect information from all hospitals for people admitted with traumatic injuries. The database is used to improve services and anonymously to help researchers answer specific research questions.

### Investigators

**Chief Investigator**
Dr Philip Braude
Consultant Geriatrician
North Bristol Trust

**Co-investigators**
Dr Mark Baxter
Consultant Orthogeriatrician
University Hospital Southampton

Dr Frances Parry
Specialist Registrar Geriatric Medicine
North Bristol Trust

Dr David Shipway
Consultant Geriatrician
North Bristol Trust

Dr Julian Thompson
Consultant Anaesthetist
North Bristol Trust
Lead for Severn Major Trauma Network

Prof Fiona Lecky
Professor of Emergency Medicine
University of Sheffield
TARN Research Director

**Study Statisticians**
Dr Omar Bouamra
Medical Statistician
University of Manchester

Dr Ben Carter
Reader in Medical Statistics
King's College London

### Analysis Plan

### 1. Background of the Study

The majority patients sustaining major trauma are 65 years or older. In March 2019 it was mandated by NHS England and NHS Improvement that a frailty assessment be performed within 72 hours of admission [1]. The Trauma and Audit Research Network (TARN) database collates data across hospitals in England and Wales admitting traumatic injuries. Patients' frailty scores are recorded on the TARN database, alongside their injury severity score, mechanism of injury, and various outcome measures.

The frailty assessment uses the Clinical Frailty Scale (CFS), which has been validated as a predictor of institutionalisation and mortality in the non-trauma healthcare setting [2]. There have been two studies in North America showing an association between CFS and outcome in major trauma patients [34]. Studies undertaken at North Bristol Trust examined the outcomes of 585 patients who were 65 years old or over admitted to the major trauma centre. They found that frailty, using the Clinical Frailty Scale, was an independent predictor of 30-day mortality [5] and 1 year mortality [6].

In a similar work looking at frailty and patient outcomes, the ELF study (Emergency Laparotomy and Frailty) [7] found that the CFS is a predictor of mortality and level of care required at discharge, and that it should be used to inform decision making and discharge planning in surgical units. Beyond this, the National Emergency Laparotomy Audit (NELA) published a report demonstrating that postoperative review by a geriatrician was associated with lower mortality [8]. We aim to look at similar outcome measures as both analysed by Rickard et al (2021), and Braude et al (2021) for the national TARN dataset.

#### *1.1 Principal research objectives to be addressed*

**Primary objectives**

To determine the association between pre-injury frailty and mortality.

**Secondary objectives**

To assess the association of pre-injury frailty:

- Inpatient mortality
- Readmission
- Post-injury complications
- Length of hospital stay

#### *1.2 Study design*

FiTR is a prospective cohort study of patients experiencing traumatic injuries admitted to the 22 major trauma centres in England. These data were collected between 1st March 2019 (from introduction of the Best Practice Tariff including frailty scoring recorded in TARN) and 31st October 2021.

**Exposure under investigation**

A requirement of achieving the BPT was that frailty was assessed by a geriatrician: a consultant, non-consultant career grade (NCCG), or specialist trainee ST3 or above. The Clinical Frailty Scale was used, which is an unstructured judgement based frailty tool using an ordinal hierarchical scale [2]. The original version 1.0 of CFS was published in 2005. The tool changed slightly towards the end of the data collection in September 2020 to Version 2.0 [9]. The fundamentals of the tool are similar enough that with scores ranging from one 'very fit' to nine 'terminally ill'.

In keeping with other studies using CFS as a prognostic tool, for the analyses, the CFS will be categorised and reported using Version 2.0 for updated terminology as: not frail (CFS 1-3), living with very mild frailty (CFS 4), living with mild frailty (CFS 5), living with moderate frailty (CFS 6), living with severe frailty (CFS 7), living with very severe frailty (CFS 8), terminally ill (CFS 9). If there are less than 50 cases of patients with CFS 8, then CFS 7 were be combined with CFS 8 and described as severely and very severely frail. CFS 9 will be reported separately.

Missing CFS will be explored, and the primary analysis will exclude these cases. However, if pattern missingness is found this may be imputed.

**Blinded Analysis Plan**

The Statistical Analysis Plan (SAP) has been drafted by the author (BC) and without prior knowledge of the data, so will be described as analyst blinded. Any changes to the primary analysis after knowledge of the outcome data will be described in a section: update of the SAP after access to the outcome assessments. Any changes to the SAP will be provided as an appendix to the SAP.

.

#### *1.3 Timing of assessments and duration of follow-up*

Inpatient mortality will be assessed. Patients will be recorded for the following outcomes at Day 30: Readmission. Post procedure complications will be recorded from the index-admission.

#### *1.4 Eligibility screening*

**Inclusion:**

1. Aged 65 years old and over
2. Admitted to a hospital within the TARN network
3. As per the TARN Standards of Practice for inclusion [10].
    a. Either:
       Direct admissions. One of:
        i. Trauma admissions whose length of stay is 3 overnight stays or more
        ii. Trauma patients admitted to a High Dependency Area regardless of length of stay
        iii. Deaths of trauma patients occurring in the hospital including the Emergency Department (even if the cause of death is medical)
        iv. Trauma patients transferred to other hospital for specialist care or for an ICU/HDU bed.

       OR
    b. Patients transferred in. One of:
        i. Trauma patients transferred into a hospital for specialist care or ICU/HDU bed whose combined hospital stay at both sites is 3 overnight stays or more
        ii. Trauma admissions to a ICU/HDU area regardless of length of stay
        iii. Trauma patients who die from their injuries (even if the cause of death is medical)

       AND
    c. Injuries meet the required criteria – See Section C in Procedures Manual [10]

**Exclusion:**

- Patients transferred in for rehabilitation should not have been submitted to TARN
- Military personnel injured on active duty should not have been submitted to TARN

#### *1.5 Data collection Measures*

What follows is a brief overview to aid understanding of the analysis plan.

**Primary outcome measure**

All-cause inpatient mortality will be assessed at the latest point recorded when the patient survival status was known. Admission to the trauma network (TN) will be coded as day 0 and the time to mortality will be measured in days to mortality. Patients that are assessed alive of the last day of follow up will be censored at this date. Any patients who die on the day of admission will be coded as 0.5 (days).

**Secondary outcome measures**

i. Day 30 readmission will be recorded as any readmission to the major trauma centre regardless of cause. If readmission to any centre in the TN is available in the dataset, this will be used in preference. Patients recorded as missing for Day 30 readmission will be assumed to be not re-admitted.
ii. Post-injury (and procedure) complications. Patients who are recorded as have any complication will be included as experiencing an event. Patients with missing complications will be imputed as not experiencing a complication.
iii. Length of stay. This will be coded as the length from time from admission to the index hospital in the TN to the date of discharge (in days). Inpatient mortality will be used as the date censored.
iv. Length of stay in critical care (as reported).

**The following clinical data will also be collected:**

i. Demographic data to include: age, sex, frailty score.
ii. Index event data to include: date of incident, date of arrival, admitting service, transferred (Y/N), type of injury, mechanism of injury intent, injury severity score, primary body area injured, pre-existing medical conditions (Charlson Comorbidity Index)
iii. Index admission procedures: surgery, complications (limited to: deep venous thrombosis, duodenal ulcer, pulmonary embolus, multiorgan failure)
iv. Involvement of a geriatrician. This will be defined by the level of the expenditure in paid clinical sessions (Direct Clinical Care (DCC)), per eligible admissions for frailty scoring, for each major trauma centre.

#### *1.6 Sample size justification*

From a single MTC study we found that one-year all-cause mortality was approximately 10% within CFS 1-3, and this increased to 20% in CFS 4-6. In order to detect this difference (Hazard Ratio = 2.12) with a 2.5% significance level (adjusted for two comparisons), and 90% power 100 events would be needed from CFS 1-6. We estimate that no more than 10% of patients will censored due to loss to follow up so will require a minimum of 800 participants and this will be inflated to 1000 by including CFS 1-8.

#### *1.7 Brief description of proposed analyses and any pre-analysis statistical checks required*

Analyses will be carried out by the study statistician following the agreed SAP. The primary analyses will use the intention-to-treat population and analysed using a mixed effects multivariable Cox regression model. Analyses will be presented as adjusted hazard ratio (aHR) with the associated 95% confidence interval and p-value. A log-log plot of the residuals will be inspected to assess the baseline proportionality assumption.

Analysis will be carried out using Stata version 16. The significance level of 5% (two-sided) will be used for all outcomes.

### 2. Data analysis plan – Data description

#### *2.1 Recruitment and representativeness of recruited participants*

CONSORT flow chart will be constructed from the full dataset to present any participant exclusion [11].

#### *2.2 Descriptive assessment of all-cause mortality*

All outcome measures listed in section 1.5 will be summarised overall, by frailty group, admission demographic, and clinical characteristics using means and standard deviation, or numbers and proportions, as appropriate. Missing data will be presented within this table.

All-cause mortality, and length of stay, will be presented graphically with a Kaplan Meier plot with associated at risk table.

#### *2.3 Loss to follow-up and other missing data*

The baseline characteristics of those missing follow-up will be compared to those with completed follow-up.

**Missing admission data**

We anticipate missing admission covariate data. The number of participants with complete data will be reported and may be imputed using a method suitable to the variable as per the recommendations of White and Thompson (14).

**Missing outcome data**

Analyses will be undertaken assuming outcomes are missing-at-random (MAR) and using all available data.

Sensitivity analyses will be carried out to assess the association of missing data.

### 3. Data analysis plan – Inferential analysis

#### *3.1 Main analysis of primary outcome*

The main analysis will use a modified intention-to-treat (ITT) population unless otherwise specified.

The primary analyses will use the intention-to-treat population and analysed using a mixed effects multivariable Cox regression model, conditioning on admission: age group (65-74, 75-84, 85+), sex, Charlson Index (0, 1, 2, 3, 4, 5+), injury severity score (9-15, 16-25, >25) [12], primary body part injured (abdomen, chest, face, head, limbs, spine, multiple, other), mechanism of trauma (fall less than 2m, fall more than 2m, vehicle incident, blows, crush, shooting & stabbing), alongside the primary exposure CFS (1-3, 4, 5, 6, 7-8). We will fit a random intercept for hospital (site). Analyses will be presented as an adjusted hazard ratio (aHR) with the associated 95% confidence interval and p-value. A log-log plot of the residuals will be inspected to assess the baseline proportionality assumption.

#### *3.2 Analysis of secondary outcomes*

**Mortality, readmission and index admission complications**

The outcome will use the intention-to-treat population and analysed using a mixed effects multivariable logistic regression model, conditioning on admission: age group (65-74, 75-84, 85+), sex, Charlson Index (0, 1, 2, 3, 4, 5+), injury severity score (9-15, 16-25, >25), primary body part injured (abdomen, chest, face, head, limbs, spine, multiple, other), mechanism of trauma (fall less than 2m, fall more than 2m, vehicle incident, blows, crush, shooting & stabbing), and CFS (1-3, 4, 5, 6, 7-8). We will fit a random intercept for hospital (site). Analyses will be presented as an adjusted odds ratio (aOR) with the associated 95% confidence interval and p-value.

**Length of hospital Stay and critical care stay**

The outcome analyses will use the intention-to-treat population and analysed using a mixed effects multivariable Cox regression model, conditioning on admission: age group (65-74, 75-84, 85+), sex, Charlson Index (0, 1, 2, 3, 4, 5+), injury severity score (9-15, 16-25, >25), primary body part injured (abdomen, chest, face, head, limbs, spine, multiple, other), mechanism of trauma (fall less than 2m, fall more than 2m, vehicle incident, blows, crush, shooting & stabbing), and CFS (1-3, 4, 5, 6, 7-8). We will fit a random intercept for hospital (site). Analyses will be presented as an adjusted hazard ratio (aHR) with the associated 95% confidence interval and p-value. A log-log plot of the residuals will be inspected to assess the baseline proportionality assumption.

#### *3.3 Populations under investigation*

**Primary outcome population – Modified Intention to Treat (ITT)**
The primary analyses will use the modified intention-to-treat (ITT) population that will include all participants eligible

#### *3.4 Sensitivity analyses*

**Non-ignorable missing outcome data**
Patients who experience mortality prior to discharge are not able to exhibit readmission during the period of investigation. Thus, patients who have died prior to discharge will excluded from the analysis

**Non-ignorable missing outcome data**
Sensitivity analyses will be used to assess the robustness of conclusions to non-ignorable missing outcome data. If this is found an inverse proportional weighting analysis will be carried out to assess the impact of the missingness.

**Transfers in**
Sensitivity analysis will be conducted after excluding all transfers in.

#### *3.5 Planned subgroup analyses*

Given that this study is not powered to detect statistically significant between-group differences, subgroup analyses are not required. We will explore the following comparisons:

i. Sex differences (male vs female)
ii. Age group
iii. Admission injury severity
iv. Primary body type injured
v. Mechanism of injury
vi. Geriatrician input

### 4. Software

Statistical analysis: Stata 16 will be used for data manipulation, description analysis and inferential analyses. R may additionally be used for production of graphs, tables, and reports.
