## Supplementary material for "A retrospective observational study to investigate the effect of frailty on outcomes of older adults admitted with major trauma": FiTR 2 Statistical Analysis Plan 0.53

**Short Title:** The FiTR 2 Study (Frailty in Trauma Reporting)

Statistical Analysis Plan
23rd December 2021
Version 0.53
IRAS: 303960

**APPROVED**
*By Philip Braude at 11:28 pm, Dec 23, 2021*

| | | |
|---|---|---|
| FiTR Study Statistician<br>Dr Ben Carter | Reader in Medical Statistics | 16th December 2021 |
| FiTR Chief Investigator<br>Dr Philip Braude | Consultant Geriatrician | 16th December 2021 |
| TARN Lead Methodologist<br>Dr Omar Bouamra | Medical Statistician | 17th December 2021 |
| TARN Research Director<br>Prof Fiona Lecky | TARN Research Lead | 23rd December 2021 |

**Authors:**

**Study Statistician**

Dr Ben Carter,

Reader in Medical Statistics,

Department of Biostatistics and Health Informatics

CLARITY (Collaborative Ageing Research) group

Institute of Psychiatry, Psychology and Neuroscience

King's College London

**Chief Investigator**

Dr Philip Braude,

Consultant Geriatrician,

CLARITY (Collaborative Ageing Research) group

REACH (Research in Emergency Care Avon Collaborative Hub)

North Bristol NHS Trust

On behalf of FiTR investigators

### Plain Language Summary

FiTR 2 study will look at older adults admitted to hospital with serious traumatic injuries across England. We plan to look at how a geriatrician review affects a person's recovery after a major injury. Geriatric assessment has been shown in single centres to reduce mortality up to a year after injury.

The information used for this study is held by the national database run by the Trauma and Audit Research Network (TARN) and is managed by the University of Manchester. TARN collect information from all hospitals for people admitted with traumatic injuries. The database is used to improve services and anonymously to help researchers answer specific research questions.

### Investigators

**Chief Investigator**
Dr Philip Braude
Consultant Geriatrician
North Bristol Trust

**Co-investigators**
Dr Mark Baxter
Consultant Orthogeriatrician
University Hospital Southampton

Dr Frances Parry
Specialist Registrar Geriatric Medicine
North Bristol Trust

Dr David Shipway
Consultant Geriatrician
North Bristol Trust

Dr Julian Thompson
Consultant Anaesthetist
North Bristol Trust
Lead for Severn Major Trauma Network

Prof Fiona Lecky
Professor of Emergency Medicine
University of Sheffield
TARN Research Director

**Study Statisticians**
Dr Omar Bouamra
Medical Statistician
University of Manchester

Dr Ben Carter
Reader in Medical Statistics
King's College London

### Analysis Plan

#### 1. Background of the Study

Older people represent the largest group of patients admitted with traumatic injuries. Fall from standing height has become the most common mechanism of injury (1). Contributing factors for this change include the proportion of older people increasing in the general population living longer with multimorbidity and associated polypharmacy, including drugs such as anticoagulation that may complicated injuries. In addition factors such as frailty, more ready access to investigations, and better awareness of the issues surrounding equity of care.

To meet the demands of this demographic change, new pathways of care and healthcare professionals trained in geriatric medicine and trauma have been developed. The core of this approach is comprehensive geriatric assessment (CGA), which is a multidomain review and management plan, delivered by a multidisciplinary team, and developed over time. There is no standard definition or composition of CGA (2). A systematic review in 2019 looking at geriatric assessment, which included some toolkits and non-geriatricians delivering care, showed a reduction in length of stay for older people with injuries, but less definitive evidence for mortality reduction (3). Cochrane have published a review examining the effect of CGA in surgical services; seven out of the eight included randomised controlled trials examined a hip fracture population. It showed long-term reduction on mortality and admission to care homes. These data could be extrapolated to the more heterogenous trauma population. A recent study looking all trauma pathology at a single centre, lead from North Bristol Trust, showed a significant reduction in mortality associated with a geriatric assessment. The study published in Annals of Surgery forms the pilot for FiTR 2.

In March 2019 NHS England and NHS Improvement added a criteria to the Best Practice Tariff (BPT) that a frailty assessment should be performed for all over 65 year olds within 72 hours of admission (4). Frailty scores were captured in the Trauma and Audit Research Network (TARN) database covering hospitals in England and Wales. Assessments were required to be delivered by a geriatrician – consultant or specialist trainee at least at year 3 (ST3+). However, this guidance did not include the need for a geriatric assessment beyond a frailty score. From communication with the Major Trauma Geriatrician Network, many trauma centres provided additional geriatric assessment and intervention beyond the financially incentivised frailty assessment. In addition, an unpublished national survey of MTCs in England indicated a wide range of funding for geriatricians to meet these targets.

FiTR 2 will explore the association between a patient receiving a frailty score, as a surrogate maker for a geriatrician review, and patient outcomes. In addition, it will examine a dose-response by exploring the association with geriatrician funded time and patient outcomes.

#### *1.1 Principal research objectives to be addressed*

**Primary objectives**

To determine the association between geriatrician review and mortality.

**Secondary objectives**

To assess the association of post-injury geriatrician review:

- Inpatient mortality
- Readmission
- Post-injury complications
- Length of hospital stay

#### *1.2 Study design*

FiTR is a prospective cohort study of patients experiencing traumatic injuries admitted to the 22 major trauma centres in England. These data were collected between 1st March 2019 (from introduction of the Best Practice Tariff including frailty scoring recorded in TARN) and 31st October 2021.

**Exposure under investigation**

A requirement of achieving the BPT was that frailty was assessed by a geriatrician: a consultant, non-consultant career grade (NCCG), or specialist trainee ST3 or above.

The exposure of geriatric assessment (GA) will be examined firstly as a binary categorisation, and secondly with a dose-response:

1) The control will be those patients eligible for to meet BPT criteria, but did not receive a frailty score, as a surrogate for no geriatrician review. The exposure will be those patients that were eligible to meet BPT criteria and had a frailty score recorded as a surrogate for geriatrician review.

   The primary coding of GA will be recorded in a binary manner. No GA [Reference] versus a CFS score indicative of a GA

2) The level of funding provided will be used to determine a dose-response of geriatrician review. Funded time is examined in the number of half-days of funded consultant time – labelled with in NHS consultant contracts as a Direct Clinical Care (DCC) session for recording within a weekly rota (e.g. 1 DCC = ½ day per week). ST3+ sessions will be examined as equivalent to consultant DCC session, given both healthcare professionals within the same service are likely delivering the similar types of intervention. These are usually recorded as Whole Time Equivalents (WTE) with 1.0 WTE being a 5-day week role (e.g.1 consultant DCC = 1 ST3+ 0.1 WTE). Across all 22 MTCs these data will be divided into quartiles using the total funded time per centre over number of patients with a frailty score.

Coding for Geriatric Assessment Score (GAS)

0 = No CFS (Reference)
1 = CFS recorded and the lowest quartile of geriatrician funding
2 = CFS recorded and the $2^{nd}$ quartile of geriatrician funding
3 = CFS recorded and the $3^{rd}$ quartile of geriatrician funding
4 = CFS recorded and highest quartile of geriatrician funding

#### *1.6 Sample size justification*

The primary analysis sample size was defined as the total records included within the TARN dataset. Using data from a single MTC we found a clear effect of GA and anticipate that this will be replicated throughout all centres (6).

#### *1.7 Brief description of proposed analyses and any pre-analysis statistical checks required*

Analyses will be carried out by the study statistician following the agreed SAP. The primary analyses will use the intention-to-treat population and analysed using a mixed effects multivariable Cox regression model. Analyses will be presented as adjusted hazard ratio (aHR) with the associated 95% confidence interval and p-value. A log-log plot of the residuals will be inspected to assess the baseline proportionality assumption.

Analysis will be carried out using Stata version 16. The significance level of 5% (two-sided) will be used for all outcomes.

### 2. Data analysis plan – Data description

#### *2.1 Recruitment and representativeness of recruited participants*

CONSORT flow chart will be constructed from the full dataset to present any participant exclusion (7).

#### *2.2 Descriptive assessment of all-cause mortality*

All outcome measures listed in section 1.5 will be summarised overall, by geriatrician review, admission demographic, and clinical characteristics using either means and standard deviation, numbers and proportions as appropriate. Missing data will be presented within this table.

All-cause mortality, and length of stay, will be presented graphically with a Kaplan Meier plot with associated at risk table.

#### *2.3 Loss to follow-up and other missing data*

The baseline characteristics of those missing follow-up will be compared to those with completed follow-up.

**Missing admission data**

We anticipate missing admission covariate data. The number of participants with complete data will be reported and may be imputed using a method suitable to the variable as per the recommendations of White and Thompson (14).

**Missing outcome data**

Analyses will be undertaken assuming outcomes are missing-at-random (MAR) and using all available data.

Sensitivity analyses will be carried out to assess the association of missing data.

### 3. Data analysis plan – Inferential analysis

#### *3.1 Main analysis of primary outcome*

The main analysis will use a modified intention-to-treat (ITT) population unless otherwise specified.

The primary analyses will use the intention-to-treat population and analysed using a mixed effects multivariable Cox regression model, conditioning on admission: age group (65-74, 75-84, 85+), sex, Charlson Index (0, 1, 2, 3, 4, 5+), injury severity score (9-15, 16-25, >25), primary body part injured (abdomen, chest, face, head, limbs, spine, multiple, other), mechanism of trauma (fall less than 2m, fall more than 2m, vehicle incident, blows, crush, shooting & stabbing), GA (None [Reference] versus presence). We will fit a random intercept for hospital (site). Analyses will be presented as an adjusted hazard ratio (aHR) with the associated 95% confidence interval and p-value. A log-log plot of the residuals will be inspected to assess the baseline proportionality assumption.

#### *3.2 Secondary analysis of primary outcome*

This will be a repeat of analysis 3.1, and GA (binary) will be replaced with the GAS (0 [Reference], versus 1, 2, 3, 4) in a dose response.

#### *3.3 Analysis of secondary outcomes*

**Mortality, readmission and index admission complications**

The outcome will use the intention-to-treat population and analysed using a mixed effects multivariable logistic regression model, conditioning on admission: age group (65-74, 75-84, 85+), sex, Charlson Index (0, 1, 2, 3, 4, 5+), injury severity score (9-15, 16-25, >25) [12], primary body part injured (abdomen, chest, face, head, limbs, spine, multiple, other), mechanism of trauma (fall less than 2m, fall more than 2m, vehicle incident, blows, crush, shooting & stabbing), GA (None [Reference] versus presence). We will fit a random intercept for hospital (site). Analyses will be presented as an adjusted odds ratio (aOR) with the associated 95% confidence interval and p-value.

**Length of hospital Stay and critical care stay**

The outcome analyses will use the intention-to-treat population and analysed using a mixed effects multivariable Cox regression model, conditioning on admission: age group (65-74, 75-84, 85+), sex, Charlson Index (0, 1, 2, 3, 4, 5+), injury severity score (9-15, 16-25, >25), primary body part injured (abdomen, chest, face, head, limbs, spine, multiple, other), mechanism of trauma (fall less than 2m, fall more than 2m, vehicle incident, blows, crush, shooting & stabbing), and frailty (CFS 1-3, 4-6, 7-9). We will fit a random intercept for hospital (site), and GA (None [Reference] versus presence). Analyses will be presented as an adjusted hazard ratio (aHR) with the associated 95% confidence interval and p-value. A log-log plot of the residuals will be inspected to assess the baseline proportionality assumption.

#### *3.4 Secondary analyses of 3.2 to 3.3*

This will be a repeat of analysis 3.1, and GA (binary) will be replaced with the GAS (0 [Reference], versus 1, 2, 3, 4) in a dose response.

#### *3.5 Populations under investigation*

**Primary outcome population – Modified Intention to Treat (ITT)**

The primary analyses will use the modified intention-to-treat (ITT) population that will include all participants eligible.

#### *3.6 Sensitivity analyses*

**Non-ignorable missing outcome data**

*Patient mortality*

Patients who experience mortality prior to discharge are not able to exhibit readmission during the period of investigation. Patients who have died prior will be excluded from the analysis

**Non-ignorable missing outcome data**

Sensitivity analyses will be used to assess the robustness of conclusions to non-ignorable missing outcome data. If this is found an inverse proportional weighting analysis will be carried out to assess the impact of the missingness.

**Transfers in**

Sensitivity analysis will be conducted after excluding all transfers in.

#### *3.7 Planned subgroup analyses*

Given that this study is not powered to detect statistically significant between-group differences, subgroup analyses are not required. We will explore the following comparisons:

i. Sex differences (male vs female)
ii. Age group
iii. Admission injury severity
iv. Primary body type injured
v. Mechanism of injury
vi. CFS frailty group

### 4. Software

Statistical analysis: Stata 16 will be used for data manipulation, description analysis and inferential analyses. R may additionally be used for production of graphs, tables, and reports.
