## Supplementary material for "A retrospective observational study to investigate the effect of frailty on outcomes of older adults admitted with major trauma": FiTR Protocol 1.0 (superceded)

#### Full Title of The Study

#### Short Study Title

The FiTR Study (Frailty in Trauma Reporting)

**APPROVED**
*By Philip Braude at 11:29 pm, Dec 23, 2021*

#### Protocol Version Number and Date

**Version** 1.0

**Date** 21st July 2021

**Update 23-12-21** See Statistical Analysis Plans FiTR 1 and 2 v0.53

##### Research Reference Numbers

**IRAS Number** 303960

**Sponsors Number** NBT5036

**Funders Number** Not Applicable

**REC reference** 21/HRA/3717

**HRA approval** 12th November 2021

#### Signature Page

The undersigned confirm that the following protocol has been agreed and accepted and that the Chief Investigator agrees to conduct the study in compliance with the approved protocol and will adhere to the principles outlined in the Declaration of Helsinki, the Sponsor's SOPs, and other regulatory requirement.

I agree to ensure that the confidential information contained in this document will not be used for any other purpose other than the evaluation or conduct of the investigation without the prior written consent of the Sponsor

I also confirm that I will make the findings of the study publicly available through publication or other dissemination tools without any unnecessary delay and that an honest accurate and transparent account of the study will be given; and that any discrepancies from the study as planned in this protocol will be explained.

**For and on behalf of the Study Sponsor:**

Signature: ..................................................................................................... Date: ....../....../......

Name (please print): .....................................................................................................

Position: .....................................................................................................

**Chief Investigator:**

Signature: ..................................................................................................... Date: 17../..5../.2021

Name: (please print):

.....Dr Philip Braude..........................................................

### Key Study Contacts

| | |
|---|---|
| Chief Investigator | Philip Braude, |
| Study Co-ordinator | Philip Braude, |
| Sponsor | North Bristol NHS Trust |
| Joint-sponsor(s)/co-sponsor(s) | N/A |
| Funder(s) | No specific funding was obtained for the delivery of this study.<br>- Philip Braude is supported by Research Capability Funding from North Bristol Research and Innovation Department, and has a Bristol Health Research Fellowship<br>- Ben Carter is supported through the NIHR Maudsley Biomedical Research Centre at the South London and Maudsley NHS Foundation Trust in partnership with King's College London |
| Key Protocol Contributors | Philip Braude,<br>Dr Frances Parry<br>Dr Ben Carter |

#### Study Summary

| | |
|---|---|
| Study Title | A retrospective observational study to investigate the effect of frailty on outcomes of older adults admitted with major trauma |
| Internal ref. no. (or short title) | The FiTR Study (Frailty in Trauma Reporting) |
| Study Design | Retrospective observational database study |
| Study Participants | Patient data held within the Trauma and Audit Research Network (TARN) database from 23 Major Trauma Centres in England |
| Planned Size of Sample (if applicable) | 800 participants required.<br>10,000 participants anticipated from these data. |
| Follow up duration (if applicable) | No follow-up |
| Planned Study Period | Data from April 2019 and March 2020 |
| Research Question/Aim(s) | 1) Is there any association between frailty and clinical outcomes in an older people admitted with serious traumatic injuries?<br>2) Is there any association between a geriatrician review and clinical outcomes in an older people admitted with serious traumatic injuries? |

#### Plain Language Summary

The majority of major injuries admitted to hospital are now in older people, with a fall from standing height being the most common reason for injury. Our study will look at older adults admitted to hospital with serious traumatic injuries across England and Wales. It will aim to work out firstly if there is an affect of frailty on peoples' survival after injury. Frailty is the reserve a person has to cope with illness and is a measure of a person slowing down over time usually due to the collection of lots of health problems. Secondly we will look at if being seen by a old age specialist (geriatrician) has an effect on a person's chances of surviving their injuries. The records we will look at are held collected routinely and held by a national database run by the Trauma and Audit Research Network (TARN) from the University of Manchester. They collect information from the 23 centres of major trauma excellence around the country and help researchers to work with them to access the anonymous data for specific research questions. We aim to use these results to help clinicians and health systems to improve how the fund and deliver care for older people.

### Funding and Support in Kind

| FUNDER(S) | FINANCIAL AND NON FINANCIALSUPPORT GIVEN |
|---|---|
| Research Capability Funding from North Bristol Research and Innovation Department. | Financial. Not directly for this study. |
| NIHR Maudsley Biomedical Research Centre at the South London and Maudsley NHS Foundation Trust in partnership with King's College London | Financial. Not directly for this study. |
| Trauma and Audit Research Network (TARN) | Non-financial personnel support |

#### *Role of Study Sponsor and Funder*

North Bristol Trust R&I - supporting the development of this work. No direct influence in any part of the work.
TARN – supporting development of the work. Will have input into the development, analysis and write-up of the work.

#### *Study Steering Group*

| | Role | Responsibility |
|---|---|---|
| Dr Philip Braude | Consultant Geriatrician North Bristol Trust | Lead for study at NBT. |
| Prof Fiona Lecky | Professor of Emergency Medicine at the University of Sheffield | Lead for study at TARN. |
| Dr Omar Bouamra | Medical Statistician University of Manchester. TARN. | Study development and delivery |
| Dr Ben Carter | Senior Lecturer in Biostatistics, King's Clinical Trials Unit.<br>Honorary Senior Lecturer North Bristol Trust | Study development and delivery |
| Dr Mark Baxter | Consultant Orthogeriatrician at University Hospital Southampton<br><br>TARN Older Peoples' Group | Study development and delivery |

| | | |
|---|---|---|
| Dr Frances Parry | Specialist Registrar Geriatric Medicine | Study development and delivery |
| Dr David Shipway | Consultant Geriatrician North Bristol Trust | Study development and delivery |
| Dr Julian Thompson | Consultant Anaesthetist North Bristol Trust.<br>Lead for trauma research Severn Network | Study development and delivery |

##### *Protocol Contributors*

| | Role | Responsibility |
|---|---|---|
| Dr Philip Braude | Consultant Geriatrician North Bristol Trust | Wrote the protocol. Writing of manuscript and dissemination |
| Dr Ben Carter | Senior Lecturer in Biostatistics, King's Clinical Trials Unit.<br>Honorary Senior Lecturer North Bristol Trust | Wrote the protocol. Analysis of data. Writing of manuscript and dissemination |
| Dr Frances Parry | Specialist Registrar Geriatric Medicine | Wrote the protocol. Writing of manuscript and dissemination |

No patients were involved in the development of the protocol.

**Key Words:** Frailty, trauma, elderly care, geriatric medicine

#### Study Flow Chart

| | May 21 | Jun 21 | Jul 21 | Aug 21 | Sep 21 | Oct 21 | Nov 21 | Dec 21 | Jan 22 |
|---|---|---|---|---|---|---|---|---|---|
| R&I sponsorship | | | | | | | | | |
| Data sharing agreement signed | | | | | | | | | |
| Data transferred | | | | | | | | | |
| Data analysed | | | | | | | | | |
| Manuscript written | | | | | | | | | |
| Manuscript published | | | | | | | | | |

#### *8.2 Research Ethics Committee (REC) and other Regulatory review*

REC approval is not required. TARN holds Health Research Authority approval (Patient Information Advisory Group (PIAG) Section 251) for research on the anonymised data it maintains from NHS Trusts. "*Section 251 of the NHS Act 2006 allows the common law duty of confidentiality to be set aside for the collection and use of patient identifiable information. Approval is only given where the work aims to improve patient care and is in the public interest.*" [TARN website]. A data sharing agreement will be completed in order for NBT to access and analyse these data.

### 10 References

1. Trauma Audit and Research Network (UK). Major trauma in older people 2017

2. NHS England and NHS Improvement. 20 National Tariff Payment System-a consultation notice annexe DrD:guidance on best practice tariffs. 2019

### 11. Appendicies

#### *11.1 Appendix 1- Required documentation*

Curriculum vitaes of the research team will be submitted

#### *11.2 Appendix 2 – Schedule of Procedures*

| | **Jul 21** | **Aug 21** | **Sep 21** | **Oct 21** | **Nov 21** | **Dec 21** | **Jan 22** | **Feb 21** | **Mar 21** |
|---|---|---|---|---|---|---|---|---|---|
| R&I sponsorship | | | | | | | | | |
| Data sharing agreement signed | | | | | | | | | |
| Data transferred | | | | | | | | | |
| Data analysed | | | | | | | | | |
| Manuscript written | | | | | | | | | |
| Manuscript submitted | | | | | | | | | |

#### *11.3 Appendix 3 – Amendment History*

| **Amendment No.** | **Protocol version no.** | **Date issued** | **Author(s) of changes** | **Details of changes made** |
|---|---|---|---|---|
